# Satellite imagery encodes features predictive of regional mortality and life expectancy

**DOI:** 10.64898/2026.05.17.26353439

**Authors:** Yasuhito Mitsuyama, Kenichi Saito, Sota Kurimoto, Shannon L Walston, Hirotaka Takita, Daiju Ueda

## Abstract

**Background:** Increasingly accessible satellite imagery provides scalable measures of the built and natural environment relevant to population health. However, whether such imagery can capture subnational variation in mortality and life expectancy remains unclear. We therefore assessed its predictive value for regional mortality and life expectancy across OECD regions.

**Methods:** We conducted an ecological, cross-sectional prediction study using 2023 data from OECD Territorial Level 3 (TL3) regions. Annual cloud-masked composites from the Harmonized Landsat and Sentinel-2 collection were processed in the Google Earth Engine, tiled at 224 × 224 pixels, and encoded with the pretrained Prithvi foundation model to derive region-level satellite embeddings. For each outcome, we trained LightGBM regressors for a country-only baseline, a satellite-only model, a combined model (country + satellite), and a final contextual model that additionally included prespecified socioeconomic and environmental covariates. Performance was evaluated using 10-fold outer cross-validation with held-out test folds; R^2^ was the primary metric.

**Results:** The analytic sample comprised 2,414 OECD TL3 regions across 38 countries, for which 939,959 satellite image tiles were processed. In paired bootstrap comparisons, adding satellite features to country indicators improved predictive performance for all outcomes, with incremental R^2^ ranging from 0.097 to 0.233. The final contextual model achieved R^2^ values of 0.78 (95% CI, 0.74–0.81) for crude mortality, 0.87 (0.84–0.89) for age-adjusted mortality, 0.86 (0.82–0.88) for infant mortality, and 0.76 (0.69–0.84) for life expectancy. In SHAP analyses, the aggregated satellite image effect consistently ranked among the top predictors across outcomes.

**Conclusion:** Satellite imagery captures subnational environmental heterogeneity relevant to regional mortality and life expectancy beyond country identity alone. Earth observation may therefore provide a scalable, complementary data source for characterizing geographic disparities in population health.

## Introduction

Mortality rates and life expectancy are core indicators of population health and fundamental targets for public health monitoring and policy.^1,2^ In most countries, these measures are derived from civil registration and vital statistics (CRVS) systems, which remain the standard source for deaths and life expectancy estimation.^3^ However, understanding geographic variation in these outcomes at subnational scale remains challenging, particularly when estimates must be stabilized over multiple years or reported across heterogeneous administrative units.^3^ As a result, there is continued interest in scalable approaches that can help characterize spatial disparities in mortality-related outcomes.

At the same time, Earth observation is entering an era of high-frequency, globally accessible measurement. The long-running Landsat program provides a decades-long open-access record of moderate-resolution Earth observations, the Sentinel-2 constellation provides open-access imagery with a nominal 5-day revisit at the equator, and commercial constellations increasingly support near-daily land imaging at meter-scale resolution.^4^ Together with cloud-based geospatial platforms such as the Google Earth Engine, these developments have made it feasible to process very large image collections reproducibly and at scale.^5^ Satellite imagery also encodes rich information about the built and natural environment, including urban form, transportation infrastructure, land use, and greenness, all of which may be relevant to population health.

Recent studies have shown that machine learning applied to satellite imagery can discern fine-grained spatial variation in socioeconomic conditions, including poverty and asset-based wealth, and can generalize within countries rather than simply separating richer from poorer nations.^6,7^ Satellite-derived representations have also been linked to intermediate health-related outcomes (e.g., neighborhood obesity prevalence^8^), supporting the premise that imagery captures contextual determinants relevant to health.^8^ In contrast, systematic evaluation of satellite imagery for mortality-related outcomes remains limited to a small number of proof-of-concept studies in restricted settings and with heterogeneous baselines.^9^ As a result, it remains unclear whether imagery provides information beyond broad national or regional differences.

To address this gap, we evaluated whether satellite imagery can predict small-area variation in mortality and life expectancy across OECD Territorial Level 3 (TL3) regions, a standardized geography designed for cross-country comparability.^10^ We prespecified a comparative modeling strategy contrasting a country-identity baseline, an imagery-only model, and a combined model, and we quantified the incremental explained variance attributable to satellite-derived features beyond country membership as the gain in cross-validated R^2^ in the combined model relative to the country-only model. Finally, we fit an additional contextual model incorporating conventional socioeconomic and environmental indicators to benchmark the relative contribution of satellite representations alongside established correlates.

## Methods

### Study design, setting, and reporting framework

We conducted an ecological, cross-sectional prediction study at the OECD Territorial Level 3 (TL3) regional level.^11^ The objective was to assess whether satellite imagery contains predictive information for small-area variation in mortality and life expectancy across OECD countries. The same candidate models, validation framework, and evaluation metrics were applied to each outcome under a prespecified comparative design. Reporting was structured according to the TRIPOD+AI guideline.^12^

### Data sources and units of analysis

#### Geographic units

The sampling frame comprised TL3 regions defined in the OECD territorial correspondence.^11^ Regions were included if complete TL3 boundary data were available at both the country and region level, permitting consistent spatial extraction and linkage to satellite imagery; regions not meeting this criterion were excluded (Figure 1). The unit of analysis for all predictors and outcomes was the TL3 region. Mortality-related outcomes and life expectancy for calendar year 2023 were obtained from the OECD Data Explorer at the TL3 level.^13^

**Figure 1.**
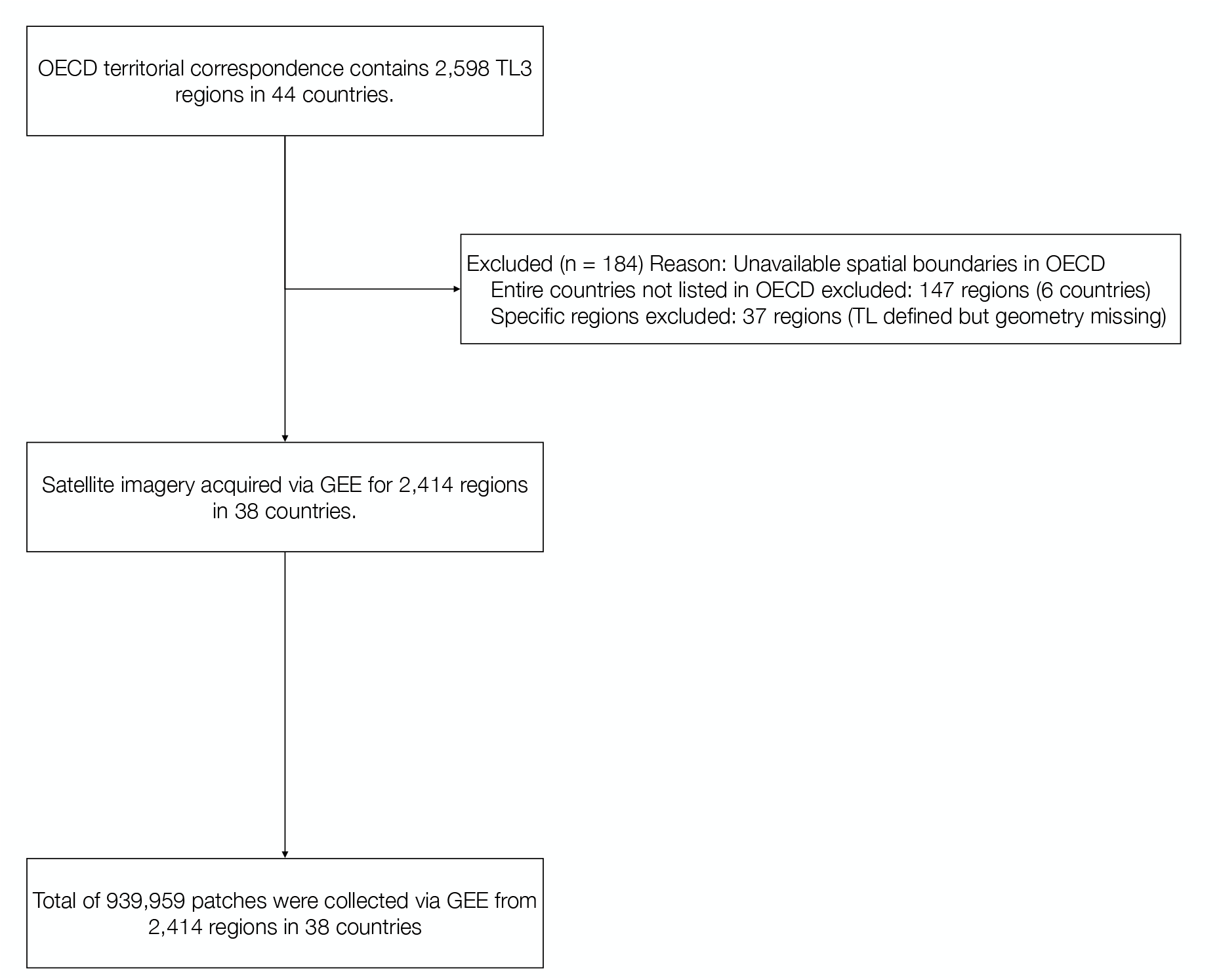
Study flow and satellite image extraction across OECD TL3 regions The OECD territorial correspondence includes 2,598 TL3 regions across 44 countries. Regions were eligible if TL3 boundaries were available for spatial processing and if satellite imagery could be linked to region geometries for the study period. After exclusions due to missing/unavailable spatial boundaries (n=184), the final analytic sample comprised 2,414 TL3 regions in 38 countries. For included regions, satellite imagery was processed in the Google Earth Engine and sampled into images, yielding a total of 939,959 images used to derive region-level satellite representations.

### Outcomes

The prespecified outcomes were TL3-level crude mortality rate, age-adjusted mortality rate, infant mortality rate, and life expectancy. All outcomes were modeled as continuous variables.

### Predictors

#### Satellite imagery and region-level representation

We accessed Harmonized Landsat and Sentinel-2 (HLS) L30 surface reflectance imagery through the Google Earth Engine for calendar year 2023.^5^ To reduce cloud-related noise and standardize temporal coverage, we generated an annual composite for each location from one year of observations. Clouds were masked using the Fmask quality band, and pixel values were aggregated with a temporal median reducer. Imagery tiles were extracted at 224 × 224 pixels and 30-m spatial resolution using six spectral bands (blue, green, red, near-infrared, SWIR1, and SWIR2), matching the HLS data structure and the input specification of the pretrained Prithvi model. Each TL3 region was represented by multiple tiles sampled across its geographic extent using a consistent extraction procedure across regions.

Rather than fitting models directly to raw pixels, we used the Prithvi (EO-1.0 / 100M) foundation model as a fixed feature extractor to generate latent image representations (embeddings) for each tile.^14^ The encoder was not fine-tuned. Because Prithvi was pretrained on HLS imagery from 2017, the 2023 imagery analyzed here did not overlap with the pretraining period. Tile-level embeddings were aggregated within each TL3 region by mean pooling to yield one satellite feature vector per region.

#### Country identity and contextual covariates

Country identity was encoded as one-hot indicators derived from the TL1 country code. These variables served both as a strong baseline predictor set and as covariates in combined models.

We additionally prespecified a set of contextual socioeconomic and environmental covariates: gross domestic product,^15^ educational attainment,^16^ unemployment rate,^17^ mean population-weighted PM2.5 exposure,^18^ population density,^19^ and dependency ratios for populations aged <15 years and ≥65 years.^20^ These variables were included only in the final contextual multivariable model to benchmark satellite-derived representations against established correlates of mortality and life expectancy.

### Missing data

Missing values in contextual covariates were imputed within each outer cross-validation training split to avoid information leakage. Specifically, missing entries were first imputed using the within-country mean (grouped by TL1) estimated from the training data; any remaining missing values were then imputed using the overall training-set mean.^21^ The resulting imputation rules were applied unchanged to the corresponding validation and test data within that fold.

### Model development and validation

#### Prespecified model set

For each outcome, we evaluated four prespecified models:

1. **Country model (baseline):** country one-hot indicators only.
2. **Satellite model:** region-level pooled satellite embeddings only.
3. **Combined model:** country indicators plus satellite features.
4. **Final contextual multivariable model:** country indicators, satellite features, and the prespecified socioeconomic/environmental covariates (Supplementary Figure S1).

To contextualize conventional determinants, we also fit univariate models for each socioeconomic/environmental covariate using the same validation framework.

#### Learning algorithm and training procedure

All models were implemented with gradient-boosted decision trees using LightGBM.^22^ We used a common training procedure across outcomes and model specifications. Early stopping was applied to reduce overfitting, with a maximum of 50 rounds without improvement on the internal validation set.

#### Cross-validation framework

We used 10-fold outer cross-validation with shuffling and a fixed random seed. Within each outer fold, the training portion was further split into a model-fitting subset and an internal validation subset used exclusively for early stopping, while the outer-fold test subset was reserved exclusively for performance evaluation. Predictions from the held-out test subsets were concatenated across the 10 folds to calculate overall performance.

#### Performance measures and incremental explained variance

The primary performance metric was the coefficient of determination (R^2^); root-mean-square error (RMSE) was reported as a complementary scale-dependent measure for the final contextual multivariable model. Incremental information attributable to satellite imagery beyond country identity was defined as the difference in R^2^ between the Combined model and the Country model.

### Model interpretation

#### Feature attribution for the Final contextual multivariable model

To characterize the relative contribution of each input domain in the final contextual multivariable model, we computed SHAP values using a tree-based explainer.^23^ Because the satellite embeddings and country indicators are high-dimensional feature groups, we summarized their overall contribution by aggregating SHAP values across all satellite embedding dimensions (“Satellite image effect”) and across all country indicator variables (“Country effect”). Socioeconomic and environmental covariates were retained as individual features in the SHAP summaries.

#### Saliency visualization for the pretrained imagery encoder

To provide qualitative insight into the image content emphasized by the pretrained encoder, we generated saliency maps for selected exemplar tiles using Integrated Gradients applied to the Prithvi model.^14,24^ Heatmaps were overlaid on the corresponding satellite images. These visualizations were intended to illustrate which image regions most influenced the encoder representation and were not interpreted as direct attributions of the downstream LightGBM predictions.

### Statistical analysis

We estimated 95% confidence intervals for R^2^ using nonparametric bootstrap resampling with 1,000 iterations. Bootstrap resampling was performed over TL3 regions using pooled held-out predictions from the 10 outer folds. To compare the Combined model with the Country and Satellite models, we used paired bootstrap resampling with identical resampled indices across models, thereby generating bootstrap distributions of performance differences. Differences were considered statistically significant when the 95% confidence interval for the paired difference excluded zero.

### Software and reproducibility

Analyses were implemented in Python using scikit-learn for cross-validation and performance metrics, LightGBM for model fitting, and SHAP for feature attribution.^23^ Satellite imagery preprocessing was conducted in the Google Earth Engine,^5^ and tile embeddings were extracted with the pretrained Prithvi model.^14^ A fixed random seed was used for cross-validation splitting and paired bootstrap resampling to support reproducibility.

## Results

### Study sample and regional characteristics

After applying the eligibility criteria, the analytic sample comprised 2,414 TL3 regions across 38 countries (Figure 1). Across these regions, 939,959 satellite image tiles were extracted and aggregated into region-level satellite features. Baseline socioeconomic and environmental characteristics and the distributions of the four prespecified primary outcomes are summarized in Table 1. Additional results for circulatory and respiratory mortality are provided in Supplementary Table 1 and Supplementary Figure 3.

**Table 1.** Demographic and health characteristics.

|  | Total | Asia | Europe | North America | Oceania | South America |
| --- | --- | --- | --- | --- | --- | --- |
| No. of countries | 38 | 4 | 26 | 4 | 2 | 2 |
| No. of TL3 regions | 2414 | 151 | 1298 | 687 | 64 | 214 |
| No. of TL2 regions | 383 | 49 | 211 | 52 | 22 | 49 |
| Total image patches | 939959 | 36266 | 136592 | 528190 | 186560 | 52351 |
| Population density | 374.16 (1116.62) | 151.53 (191.06) | 391.04 (1038.46) | 393.40 (1098.11) | 73.86 (122.07) | 409.20 (1702.43) |
| GDP per capita | 44602.14<br>(20237.48) | 43663.17<br>(9765.52) | 46161.48<br>(20172.47) | 43077.02<br>(19360.71) | 59091.52<br>(12894.13) | 37070.02<br>(23422.12) |
| Unemployment rate | 5.60 (3.27) | 7.98 (3.95) | 4.92 (3.30) | 5.90 (2.75) | 3.83 (0.44) | 7.56 (3.16) |
| Educational attainment | 37.39 (14.72) | 35.65 (8.73) | 31.46 (11.19) | 50.16 (15.82) | 34.80 (1.50) | 38.28 (8.77) |
| Dependency ratio (<15) | 25.32 (6.55) | 21.29 (2.75) | 22.41 (3.52) | 28.77 (7.15) | 26.81 (2.27) | 30.48 (8.61) |
| Dependency ratio (65≤) | 33.29 (11.31) | 35.55 (8.92) | 36.95 (8.63) | 28.25 (12.76) | 27.90 (4.66) | 29.84 (13.54) |
| PM 2.5 exposure | 10.95 (2.07) | 10.33 (2.04) | 11.28 (2.48) | 10.84 (1.08) | 10.41 (1.29) | 10.12 (1.67) |
| Crude mortality rate | 10.68 (3.17) | 10.44 (1.86) | 11.28 (3.27) | 8.48 (2.56) | 9.13 (1.93) | 10.54 (2.51) |
| Age-adjusted mortality rate | 9.45 (2.70) | 8.50 (1.22) | 10.78 (2.39) | 7.80 (1.89) | 5.19 (3.13) | 8.70 (1.36) |
| Infant mortality rate | 4.88 (3.68) | 2.91 (1.42) | 5.11 (4.05) | 5.84 (3.43) | 3.18 (0.94) | 3.25 (1.34) |
| Life expectancy | 79.89 (3.38) | 82.19 (2.03) | 79.55 (3.07) | 77.89 (5.45) | 81.76 (0.32) | 82.08 (1.71) |
| Respiratory diseases mortality rate | 83.59 (36.23) | 101.34 (32.06) | 69.83 (30.44) | 101.01 (36.27) | - | 60.94 (20.99) |
| Circulatory diseases mortality rate | 262.69 (130.98) | 308.89 (106.39) | 271.03 (155.89) | 219.10 (71.13) | - | 419.47 (73.41) |

GDP; Gross domestic product, PM; Particulate matter

### Primary analyses

Held-out test-fold performance for the prespecified model comparisons is summarized in Table 2, with fold-wise distributions shown in Figure 2.

**Table 2.**
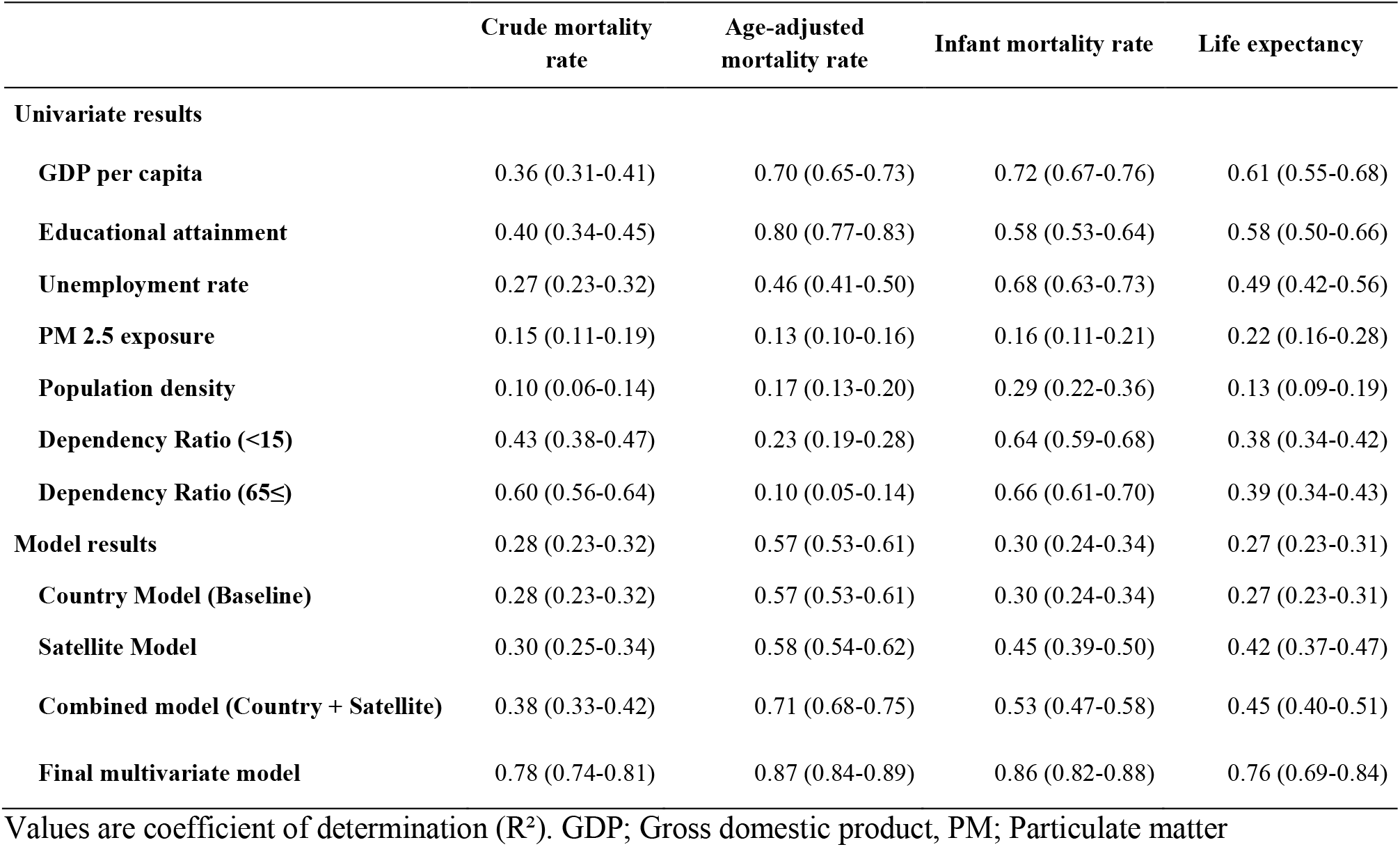
Univariate and Multivariate Analysis of Predictive Accuracy.

**Figure 2.**
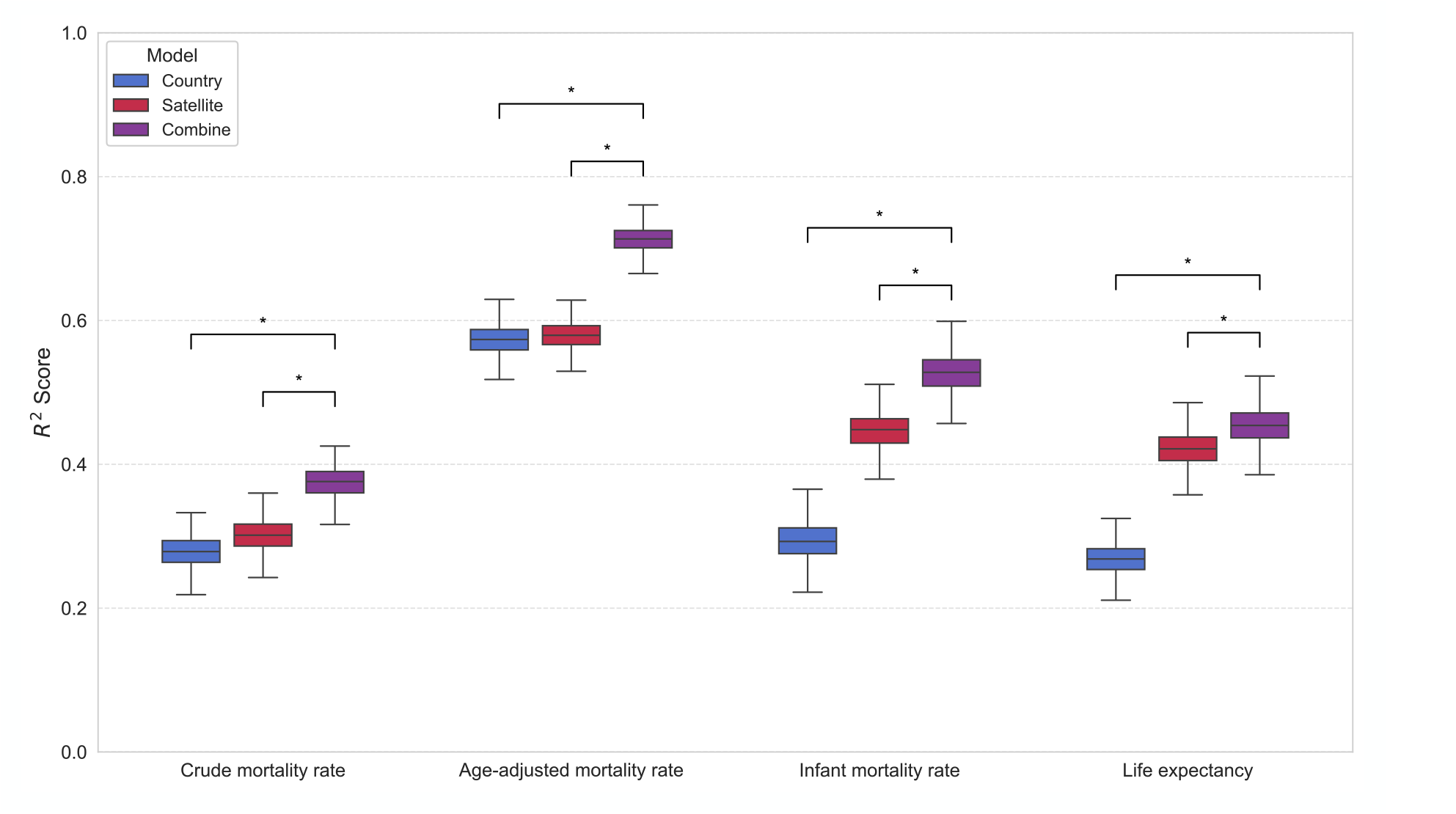
Cross-validated predictive performance of country, satellite, and combined models Boxplots show the distribution of held-out test-fold performance (R^2^) across the 10 outer cross-validation folds for three prespecified models: Country-only baseline (one-hot country indicators), Satellite-only (region-level pooled satellite embeddings), and Combined (country indicators + satellite embeddings). Performance is shown for four outcomes: crude mortality rate, age-adjusted mortality rate, infant mortality rate, and life expectancy. Brackets with asterisks denote statistically significant differences in performance between compared models based on paired resampling of held-out predictions (95% confidence interval for the difference excluding zero).

#### Country model (baseline)

Using country identity alone, the baseline model achieved R^2^ values of 0.28 (95% CI, 0.23–0.32) for crude mortality, 0.57 (95% CI, 0.53–0.61) for age-adjusted mortality, 0.30 (95% CI, 0.24–0.34) for infant mortality, and 0.27 (95% CI, 0.23–0.31) for life expectancy (Table 2).

#### Satellite model

Using satellite-derived features alone, the Satellite model achieved R^2^ values of 0.30 (95% CI, 0.25– 0.34) for crude mortality, 0.58 (95% CI, 0.54–0.62) for age-adjusted mortality, 0.45 (95% CI, 0.39– 0.50) for infant mortality, and 0.42 (95% CI, 0.37–0.47) for life expectancy (Table 2). The largest gains over the country-only baseline were observed for infant mortality and life expectancy, suggesting that satellite-derived signals captured substantial between-region variability not explained by country membership alone.

#### Combined model and incremental explained variance

Adding satellite-derived features to the country-only baseline improved performance to R^2^ values of 0.38 (95% CI, 0.33–0.42) for crude mortality, 0.71 (95% CI, 0.68–0.75) for age-adjusted mortality, 0.53 (95% CI, 0.47–0.58) for infant mortality, and 0.45 (95% CI, 0.40–0.51) for life expectancy (Table 2; Figure 2). In paired bootstrap comparisons, the Combined model outperformed the Country model for all outcomes, with mean R^2^ differences of 0.097 (95% CI, 0.058–0.135), 0.140 (95% CI, 0.110–0.169), 0.233 (95% CI, 0.183–0.282), and 0.185 (95% CI, 0.139–0.229), respectively. The Combined model also outperformed the Satellite model for all outcomes, with mean R^2^ differences of 0.073 (95% CI, 0.048–0.101), 0.133 (95% CI, 0.109–0.159), 0.080 (95% CI, 0.046–0.113), and 0.033 (95% CI, 0.018–0.050), respectively. All reported differences were statistically significant (p < 0.001). Overall, these results indicate that satellite features contributed additional explanatory information beyond country-level differences.

### Final contextual multivariable model

In the prespecified Final contextual multivariable model that incorporated satellite features, country identity, and socioeconomic/environmental indicators, predictive performance increased to R^2^ values of 0.78 (95% CI, 0.74–0.81) for crude mortality, 0.87 (95% CI, 0.84–0.89) for age-adjusted mortality, 0.86 (95% CI, 0.82–0.88) for infant mortality, and 0.76 (95% CI, 0.69–0.84) for life expectancy, with corresponding RMSE values shown in Figure 3.

**Figure 3.**
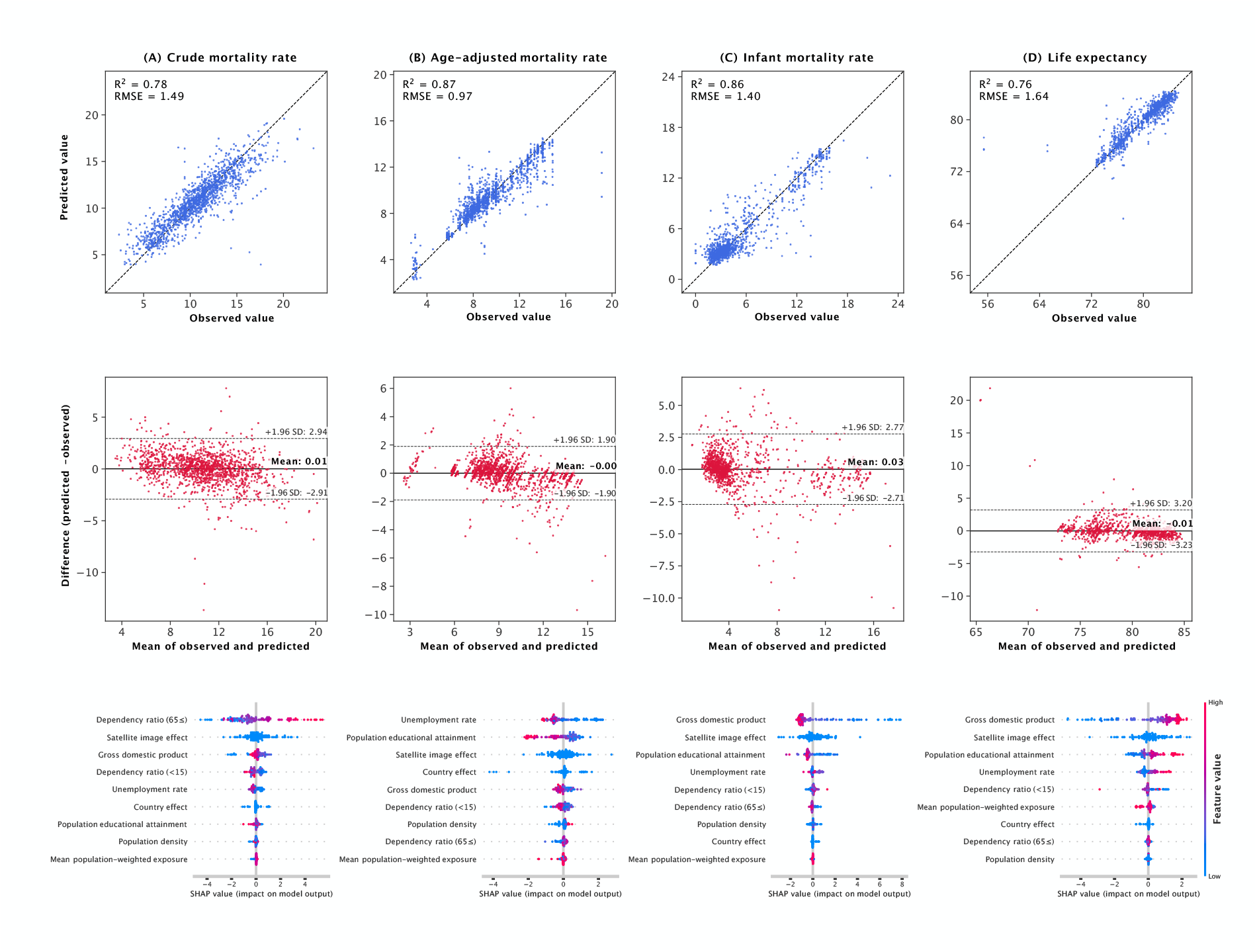
Accuracy, calibration, and feature contributions in the final contextual multivariable model Model diagnostics and interpretation for the prespecified final contextual multivariable model (country indicators + satellite embeddings + socioeconomic/environmental covariates) evaluated using held-out predictions from 10-fold outer cross-validation. (A–D) correspond to crude mortality rate, age-adjusted mortality rate, infant mortality rate, and life expectancy, respectively. **Top row:** observed vs predicted values with the identity line; inset reports overall held-out R^2^ and RMSE. **Middle row:** Bland–Altman plots showing prediction error (predicted − observed) vs the mean of observed and predicted values; horizontal lines indicate the mean difference and ±1.96 SD limits of agreement. **Bottom row:** SHAP summary plots for each outcome showing the distribution of feature impacts; “Satellite image effect” aggregates SHAP values across all embedding dimensions and “Country effect” aggregates across all country indicator variables, while socioeconomic/environmental covariates are shown individually.

### Model interpretation: SHAP summaries and qualitative saliency examples

SHAP analyses of the final contextual multivariable model showed that the aggregated “Satellite image effect” consistently ranked among the most influential predictors: second for crude mortality, infant mortality, and life expectancy, and third for age-adjusted mortality. This placed the satellite feature group alongside country identity and conventional determinants such as age structure, gross domestic product, and unemployment rate (Figure 3). As qualitative examples, saliency heatmaps overlaid on selected satellite images are presented in Supplementary Figure S4; visual inspection suggests that the encoder emphasized features such as rivers, green space, and built-up areas.

## Discussion

In this cross-sectional ecological study of 2,414 OECD TL3 regions across 38 countries, using 939,959 satellite image patches, we found that satellite imagery provided additional, quantifiable predictive information on subnational variation in mortality and life expectancy beyond country identity. The central contribution of this study is the empirical demonstration that image-derived representations can capture health-relevant regional heterogeneity at a standardized cross-national scale. The performance gains observed in the combined models provide an empirical quantification of that added information. In the final contextual models, satellite-derived effects remained among the most influential predictors.

Prior work has shown that machine learning applied to satellite imagery can recover fine-grained socioeconomic variation within countries and can be associated with intermediate health-related outcomes.^6–8^ By contrast, mortality-focused applications have remained limited to a small number of proof-of-concept studies performed in restricted settings with heterogeneous comparators.^9^ Our study extends this literature by providing a standardized cross-country evaluation at the TL3 level and by explicitly separating national context from subnational signals through a prespecified comparison of country-only, satellite-only, and combined models. This design addresses the concern that image-based models may simply encode broad country differences rather than meaningful within-country variation.

A plausible explanation for the incremental value of satellite imagery is that learned embeddings summarize complex aspects of the built and natural environment—settlement morphology, transportation networks, land-use patterns, and vegetative cover—that are difficult to capture with any single conventional indicator and may lie upstream of multiple determinants of health.^9,25–27^ Consistent with this interpretation, the largest gains from satellite features were observed for infant mortality and life expectancy, outcomes that may be especially sensitive to local living conditions and environmental context.^6,7,28,29^ In the final contextual model, aggregated SHAP values suggested that the satellite domain contributed at a level comparable to several established predictors, supporting the interpretation that satellite imagery captures health-relevant contextual variation while leaving the underlying pathways to be clarified in future work.

From a public health perspective, these findings suggest that Earth observation offers a scalable and standardized source of contextual information for monitoring geographic disparities in mortality-related indicators. CRVS systems remain the gold standard, but delays in reporting and coarse spatial aggregation can hinder timely small-area assessment^1–3^ Satellite imagery, particularly when processed through reproducible embedding pipelines, could therefore complement official statistics. At the same time, frequent image acquisition should not be conflated with real-time outcome measurement: the mortality outcomes analyzed here were not observed contemporaneously with daily or weekly resolution, so any operational use would require explicit lag structures, prospective validation, and transparent uncertainty quantification.

This study has several limitations. First, the analysis was ecological and cross-sectional, so the observed relationships should not be interpreted causally and remain susceptible to ecological fallacy.^30,31^ Second, the final contextual multivariable model incorporated contemporaneous socioeconomic and environmental indicators; its strong explanatory performance should therefore not be interpreted as evidence of prospective forecasting. Third, inclusion required administrative boundaries suitable for spatial processing, which may have introduced selection bias if excluded regions differed systematically from included regions. Fourth, generalizability beyond OECD settings remains uncertain, particularly in lower-income contexts where environmental–health relationships, data quality, and deployment risks may differ. Finally, SHAP summaries and saliency maps provide descriptive insight into model behavior but do not establish underlying mechanisms.^23,24^

In conclusion, satellite imagery contained meaningful predictive information for TL3-level mortality and life expectancy and provided incremental explanatory value beyond country identity across a broad set of OECD regions. These findings support Earth observation as a complementary data stream for characterizing subnational heterogeneity in population health. Future work should emphasize external validation beyond OECD settings, temporally explicit prediction with lagged designs, spatially structured evaluation, and robust uncertainty quantification before imagery-based models are considered for decision-oriented use.

## Data Availability

The model codes used in this study are available at https://huggingface.co/ibm-nasa-geospatial/Prithvi-EO-1.0-100M. Satellite imagery data were obtained from the Harmonized Landsat and Sentinel-2 (HLS) L30 collection through the Google Earth Engine, and mortality and life expectancy data were obtained from the OECD Data Explorer. These source datasets are publicly available from the respective providers.

## Acknowledgement

English language editing was performed using ChatGPT (GPT-5; OpenAI, San Francisco, CA, USA). All content was reviewed and validated by the authors.

