## Supplemental Appendix for "Satellite imagery encodes features predictive of regional mortality and life expectancy"

### Title

### Short title

Satellite imagery and regional mortality

### Article type

Original Article

### Supplementary Appendix

#### Supplementary Tables

**Table S1:** Univariate and Multivariate Analysis of Predictive Accuracy for Circulatory and Respiratory mortality rates

#### Supplementary Figures

**Figure S1:** Model overview

**Figure S2:** Cross-validated predictive performance of country, satellite, and combined models for circulatory disease and respiratory disease mortality rates

**Figure S3:** Accuracy, calibration, and feature contributions in the final contextual multivariable model for circulatory disease and respiratory disease mortality rates

**Figure S4:** Saliency maps highlight environmental features emphasized by the satellite encoder

#### Supplementary Tables

**Table S1:** Univariate and Multivariate Analysis of Predictive Accuracy for Circulatory and Respiratory mortality rates

| | Circulatory system mortality rate | Respiratory system mortality rate |
|---|---|---|
| **Univariate results** | | |
| **Gross Domestic Product (GDP)** | 0.24 (0.16-0.32) | 0.41 (0.35-0.48) |
| **Educational Attainment** | 0.33 (0.22-0.43) | 0.48 (0.40-0.55) |
| **Unemployment Rate** | 0.32 (0.24-0.40) | 0.38 (0.31-0.44) |
| **PM2.5 Exposure** | 0.18 (0.08-0.26) | 0.32 (0.24-0.39) |
| **Population density** | 0.06 (-0.01-0.12) | 0.41 (0.32-0.49) |
| **Dependency Ratio (<15)** | 0.39 (0.33-0.45) | 0.28 (0.22-0.34) |
| **Dependency Ratio (65≤)** | 0.47 (0.41-0.53) | 0.27 (0.21-0.33) |
| **Country Model (Baseline)** | 0.25 (0.19-0.31) | 0.43 (0.36-0.48) |
| **Satellite Model** | 0.36 (0.27-0.44) | 0.35 (0.28-0.42) |
| **Multivariate results** | | |
| **Combine model (Country + Satellite)** | 0.35 (0.26-0.44) | 0.43 (0.36-0.50) |
| **Final multivariate model** | 0.69 (0.64-0.74) | 0.69 (0.64-0.73) |

### Supplementary Figures

**Figure S1:** Model overview

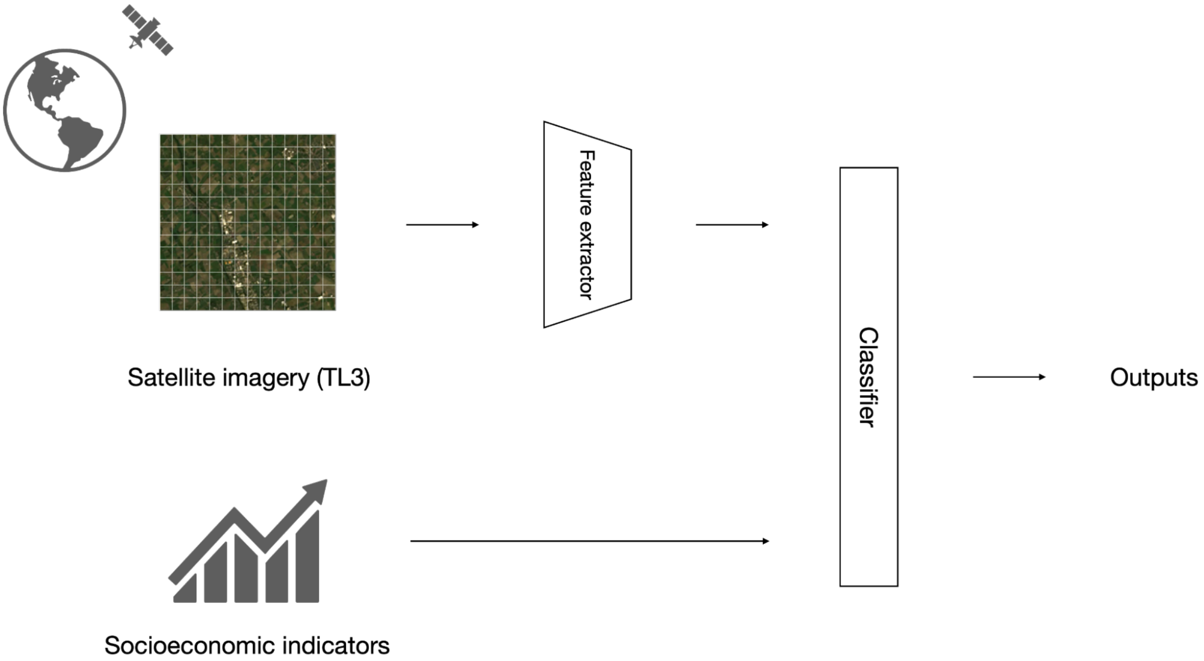


This figure outlines the deep learning model for estimating the prespecified outcomes. Fixed-length latent representations ("embeddings") were computed from each imagery tile by the Phrithvi (EO-1.0 / 100M) model as a pretrained feature extractor.[14] The classifier applied gradient-boosted decision trees (LightGBM regressor) for all analyses.[22]

**Figure S2:** Cross-validated predictive performance of country, satellite, and combined models for circulatory disease and respiratory disease mortality rates

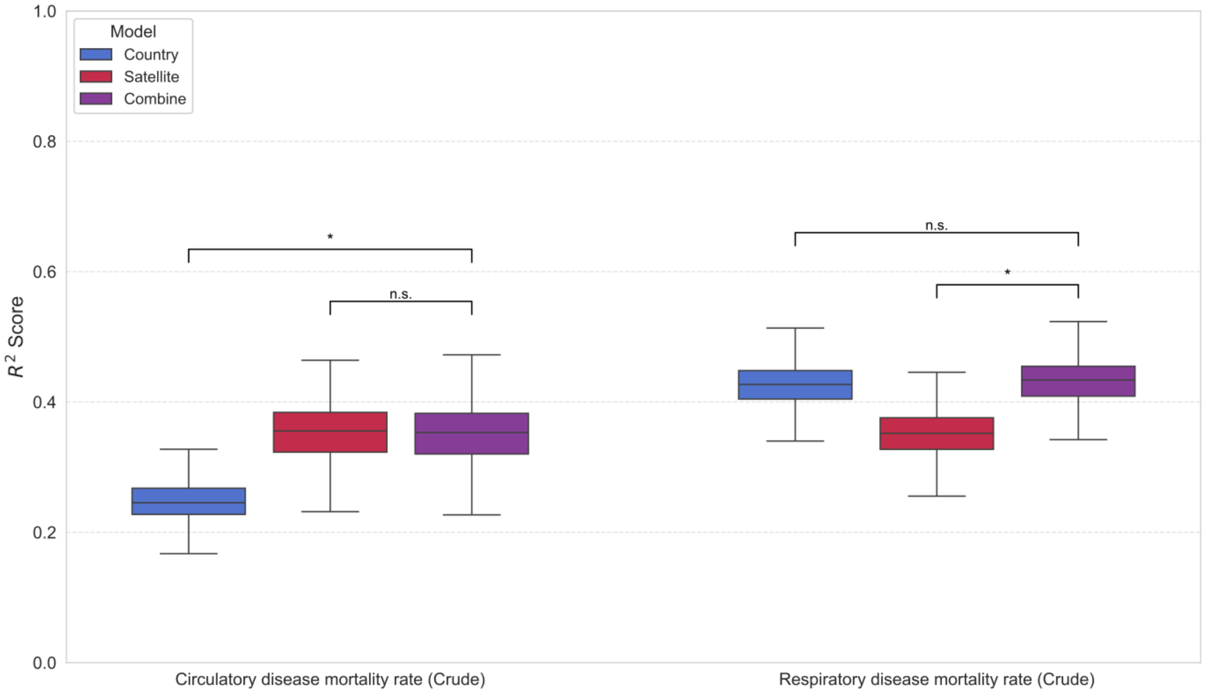


Boxplots show the distribution of held-out test-fold performance ($R^2$) across the 10 outer cross-validation folds for three prespecified models: Country-only baseline (one-hot country indicators), Satellite-only (region-level pooled satellite embeddings), and Combined (country indicators + satellite embeddings). Performance is shown for four outcomes: crude mortality rate, age-adjusted mortality rate, infant mortality rate, and life expectancy. Brackets with asterisks denote statistically significant differences in performance between compared models based on paired resampling of held-out predictions (95% confidence interval for the difference excluding zero).

**Figure S3:** Accuracy, calibration, and feature contributions in the final contextual multivariable model for circulatory disease and respiratory disease mortality rates

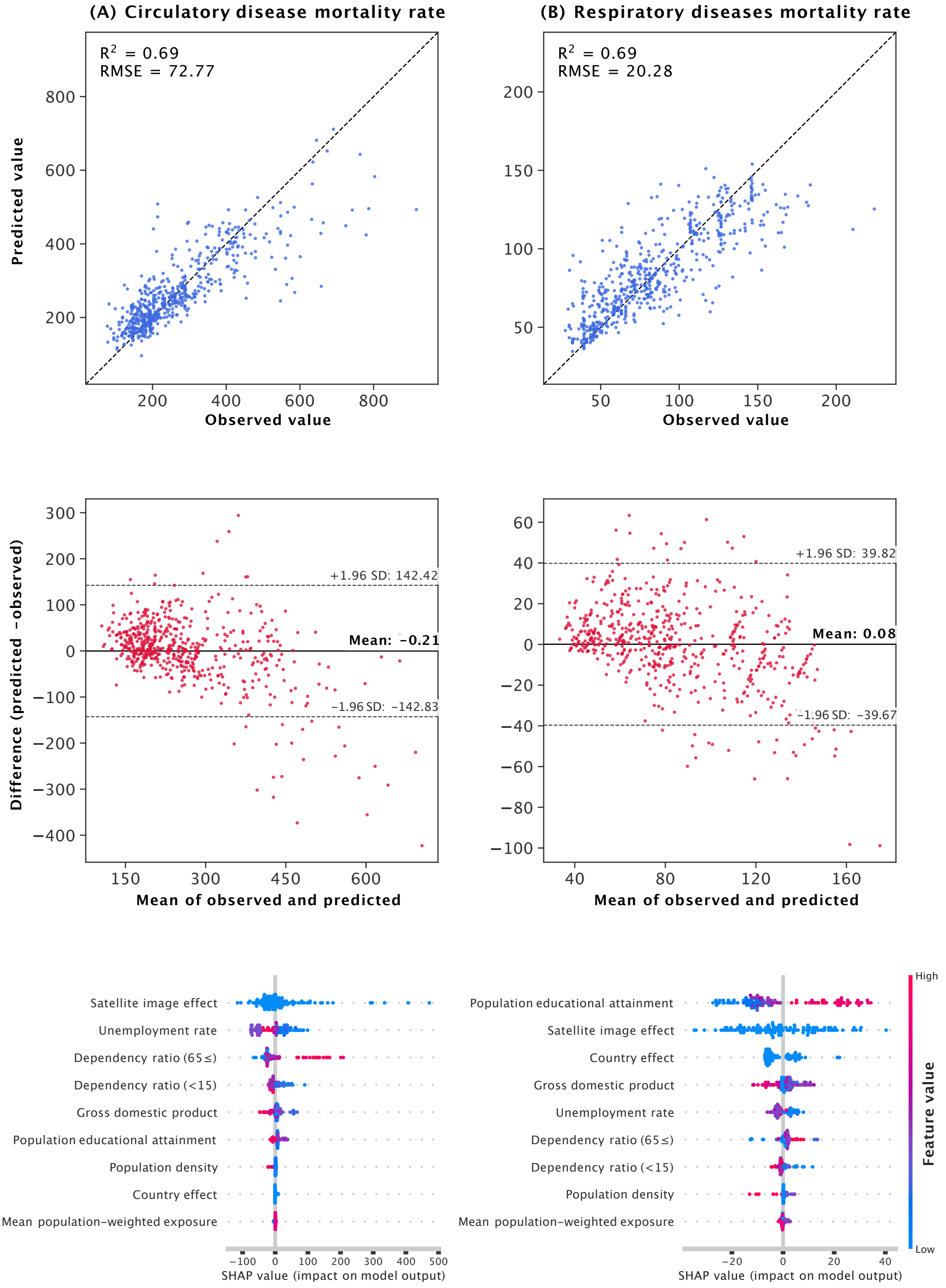


Model diagnostics and interpretation for the prespecified final contextual multivariable model (country indicators + satellite embeddings + socioeconomic/environmental covariates) evaluated using held-out predictions from 10-fold outer cross-validation.

(A) crude mortality rate due to circulatory diseases.

(B) crude mortality rate due to respiratory diseases.

**Top row:** observed vs predicted values with the identity line; inset reports overall held-out $R^2$ and RMSE. **Middle row:** Bland–Altman plots showing prediction error (predicted − observed) vs the mean of observed and predicted values; horizontal lines indicate the mean difference and ±1.96 SD limits of agreement. **Bottom row:** SHAP summary plots for each outcome showing the distribution of feature impacts; "Satellite image effect" aggregates SHAP values across all embedding dimensions and "Country effect" aggregates across all country indicator variables, while socioeconomic/environmental covariates are shown individually.

**Figure S4:** Saliency maps highlight environmental features emphasized by the satellite encoder

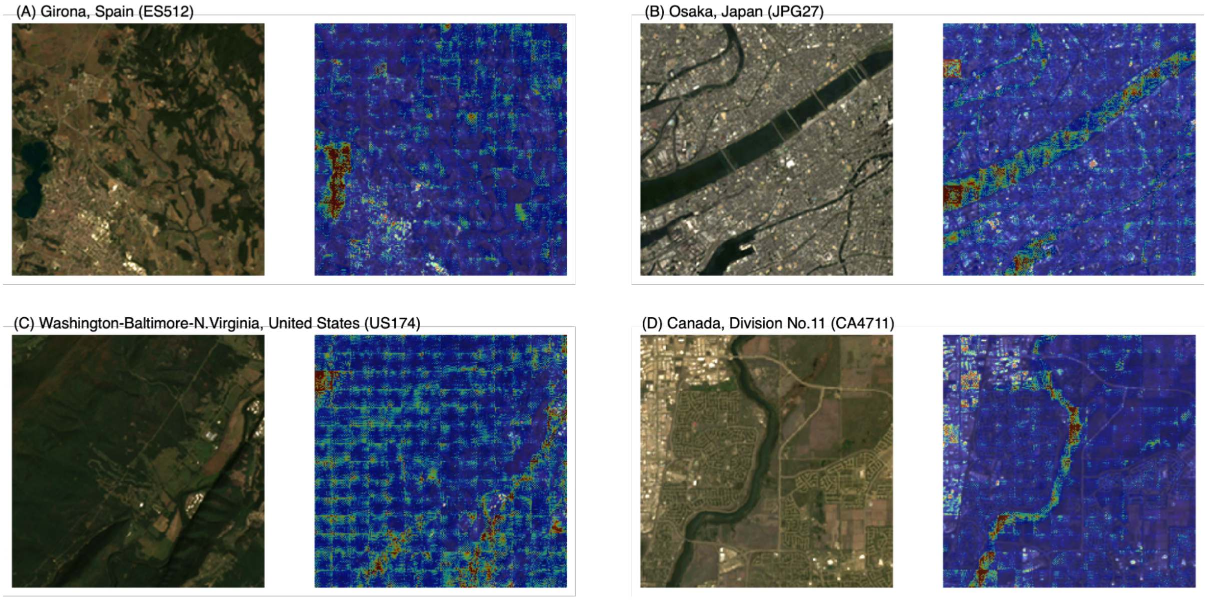


The figure presents satellite imagery (left) and corresponding saliency maps (right) for four distinct regions. (A) In Girona, Spain, the model highlights lakes, towns, and forests. (B) In Osaka, Japan, rivers and towns are emphasized. (C) In Washington-Baltimore-N. Virginia, USA, the focus is on forests and towns. (D) In Canada, Division No. 11, rivers and towns are highlighted.
